# Closing the Pneumococcal Conjugate Vaccine (PCV) Introduction Gap: An Archetype Analysis of ‘last-mile’ countries

**DOI:** 10.1101/2023.05.10.23289791

**Authors:** Preetika Banerjee, Jasmine Huber, Veronica Denti, Molly Sauer, Rose Weeks, Baldeep K. Dhaliwal, Anita Shet

## Abstract

Pneumonia remains the leading infectious cause of global childhood deaths, despite the availability of World Health Organization (WHO)-prequalified pneumococcal conjugate vaccine (PCV) products and the evidence of their safety and efficacy for over two decades, along with financial and technical support from Gavi The Vaccine Alliance (Gavi). There are 39 remaining “last-mile” countries (33 low- and middle-income countries [LMICs] and six high-income countries) that haven’t fully included PCV in their National Immunization Programs. To address this inequitable distribution of PCV, we conducted a rapid assessment and landscaping exercise of country indicators related to barriers and facilitators for PCV decision-making, aiming to categorize countries into archetypes that could benefit from shared advocacy approaches. The archetype analysis first created a country matrix focused on three domains - health characteristics, immunization factors, and policy framework - and identified ten related indicators. Countries were scored based on indicator performance and subsequently ranked and grouped into three overarching archetypes of low-, moderate-, and high-barrier to PCV introduction.

15 countries were classified as “low-barrier,” indicating that they have more factors favorable for PCV introduction, such as high immunization coverage of common childhood vaccines, recent “new” vaccine introductions, and supportive governments, as well as substantial disease burden and eligibility for Gavi support. Most of the countries classified in the “moderate-barrier” (12 countries) and “high-barrier” (6 countries) archetypes have strong immunization systems, but competing country priorities and cost barriers impede policy decision-making on PCV introduction. Other countries require strengthening of their health systems despite political will.

The barrier-based categorization can provide an actionable framework to design tailored PCV advocacy that addresses obstacles to new vaccine introductions within these “last-mile” countries. Implementation approaches that emerge from this framework can lead to strengthened decision-making on vaccine introduction and mobilization of investments in vaccine access that can enhance child survival worldwide.

## Introduction

Pneumonia (or lower respiratory infections) continues to be the leading infectious cause of death in children worldwide, accounting for 14% of deaths among children under five years (1). Preventing pneumococcal disease, a major bacterial cause of life-threatening pneumonia, sepsis, and meningitis, is vital for reducing worldwide childhood mortality while synergistically accelerating progress toward equity, education, and workforce development targets included in the United Nations (UN) Sustainable Development Goals (SDGs) (2–4).

Pneumococcal conjugate vaccines (PCV) have been used in high-income countries since 2000. Results from a multi-country analysis indicate that PCV is cost-effective in most settings and can substantially reduce the burden of disease due to *Streptococcus pneumoniae* (5,6). Mathematical modelling conducted by the Vaccine Impact Modelling Consortium (VIMC) estimates that vaccination activities for PCV between 2000-2030 could lead to approximately 2.8 million averted deaths and 190 million averted disability-adjusted life years (DALYs) (7). Currently, there are three World Health Organization (WHO) prequalified PCV products available, in five presentations that include multi-dose vials, with several other products in the pipeline. The Immunization Agenda 2030 (IA2030) aims to achieve 90% coverage for PCV by 2030 (8). However, global coverage of the final dose of PCV was only 51% in 2021 (9), with vaccine introductions and coverage having slowed down or plateaued during the COVID-19 pandemic (10).

As of January 2023, there were 39 countries that have yet to introduce PCV into their National Immunization Programs; among these countries, 33 are classified as low- and middle-income countries (LMICs) (11). Three countries (Indonesia, Tajikistan and Timor-Leste) recently introduced PCV between November 2022 and January 2023 (12–14). Barriers to vaccine introduction are specific to individual country contexts, however common barriers include sustainable financing, perception of importance and competing health priorities, human resource availability, vaccine delivery infrastructure, and functioning surveillance systems (15,16). Successful PCV introduction in these countries requires support for in-country logistical and financial planning activities along with sustained commitment from country governments, technical agencies, and donors (17).

To enhance our understanding of the current landscape and opportunities for PCV introduction, our team critically assessed key domains and related indicators that influence decision-making and implementation processes associated with PCV introduction. We aimed to highlight factors influencing country decision-making for PCV introduction and categorize countries into archetypes based on their probability of PCV introduction in the near or distant future. This type of archetypal classification, which has been used to inform policy decision-making about adult immunization, was conducted to facilitate the development of strategic and context-specific methods to address PCV introduction barriers (18). This analysis does not replace the need for strategies tailored to each country; rather, it seeks to provide meaningful insight for the global community to bridge existing gaps to significantly expand global access to PCV.

## Methods

### Country selection

Countries yet to introduce PCV (called ‘non-PCV countries’) were identified from the WHO’s Immunization, Vaccines and Biologicals (IVB) immunization database, and countries’ income level was determined using the World Bank’s country income classification (19,20). Among 194 WHO Member States, 39 countries had yet to introduce PCV into their National Immunization Programs, per WHO’s immunization data portal using 2021 data (19,21). Six high-income countries that have not yet introduced PCV – Antigua and Barbuda, Brunei Darussalam, Cook Islands, Czechia, Estonia, and Saint Kitts and Nevis – were excluded from our analysis, given the study’s focus on LMICs. The remaining geographically diverse 33 non-PCV countries with differing determinants of PCV introduction decision-making were included in the archetypal analysis.

### Mapping of domains and indicators related to vaccine introduction

Three team members independently conducted web-based searches for domains and indicators. We scanned both peer-reviewed and grey literature, media articles, country reports and guidelines, policy briefs, and resources detailing disease burden and vaccine introduction status databases from government, non-governmental sources, and from agencies such as WHO and the United Nations Child Fund (UNICEF) (Supplementary Table S1). Data were abstracted from websites that had information in English. The list of selected countries and each corresponding domain and their respective indicators were maintained on an Excel spreadsheet. All data cited for this analysis are per March 2023.

We chose domains and respective indicators from published literature pertaining to vaccine introduction. These were examined for the 33 LMICs included in this analysis (21). The three domains were chosen from an extensive list of decision-making indicators and included health characteristics, immunization factors and policy framework. The ten indicators within these domains were selected based on their likelihood to influence PCV introduction decision-making, and included 1) pneumonia incidence rate; 2) under five child mortality; 3) national health expenditure as a percentage of national gross domestic product [health domain]; 4) 2021 vaccine coverage – three doses of diphtheria, tetanus and pertussis (DTP3), 5); vaccine coverage – one dose of measles-containing vaccine (MCV1); 6) new vaccine introductions consisting of rotavirus vaccine (RV), human papillomavirus vaccine (HPV), inactivated poliovirus vaccine (IPV), or the second dose of measles-containing vaccine (MCV2); 7) partial PCV introduction status;[immunization domain]; 8) presence of a National Immunization Technical Advisory Group (NITAG); 9) political will pertaining to PCV introduction; and 10) Gavi eligibility and support [policy domain].

### Scoring framework

Each country was assigned a score between 0 to 4, corresponding to a sliding scale relevant to each indicator with its domain; the higher scores indicating stronger evidence for PCV introduction need, value, and readiness, which can lead to favorable conditions for PCV introduction (Table 2).

**Table 1:** Global Pneumococcal Conjugate Vaccine (PCV) Introduction Status as of March 2023

|  | Number of Countries |
| --- | --- |
| **<u>Total countries</u>** | **<u>194</u>** |
| Introduced PCV into National Immunization Program* | **155 (80%)** |
| Not introduced PCV into National Immunization Program<br>(including partial introduction for risk groups and/or certain regions of the country) | **39 (20%)** |
| High-income countries | 6 |
| Upper-middle-income countries | 19 |
| Lower-middle-income countries | 7 |
| Low-income countries | 6 |
| Temporarily unclassified by World Bank (Venezuela) | 1 |

\* Including Indonesia, Tajikistan and Timor-Leste who introduced PCV into their National Immunization Programs between November 2022 and January 2023.

**Table 2:** Domains, indicators and scoring framework for country archetype classification related to PCV introduction decision-making

| Domains | Indicators | Score<sup>a</sup> |  |  |  |  |
| --- | --- | --- | --- | --- | --- | --- |
|  |  | 0 | 1 | 2 | 3 | 4 |
| **Health characteristics** | 1) Pneumococcal incidence rate (2015) (cases per 100,00 children)<sup>b</sup> | <199 | 200 to 1,000 | 1,001 to 2,000 | 2,001 to 3,000 | >3,001 |
|  | 2) Child mortality rate (U5 child deaths per 1,000 live births)<sup>c</sup> | <10 | 11 to 39 | 40 to 69 | 70 to 99 | >100 |
|  | 3) National health expenditure (% of national gross domestic product - GDP) | <2.5 | 2.6-5.0 | 5.1-7.5 | 7.6-10.0 | >10.1 |
| **Immunization systems** | 4) Vaccine coverage: DTP3 (%) | less than 25% | 26-50% | 51-74% | 75 to 89% | >90% |
|  | 5) Vaccine coverage: MCV1 (%) | less than 25% | 26-50% | 51-74% | 75 to 89% | >90% |
|  | 6) Current usage of 'new' vaccines (RV, HPV, IPV, MCV2) | None | 1 vaccine introduced | 2 vaccines introduced | 3 vaccines introduced | 4 vaccines introduced |
|  | 7) PCV partial introduction | PCV not introduced (including available only in the private sector) | PCV was introduced previously but currently suspended | PCV introduced to risk groups and/or certain regions in the country (subnational introduction) | - | - |
| **Policy framework** | 8) Presence of a National Immunization Technical Advisory Group (NITAG)<sup>d</sup> | No NITAG or no information | Yes: Interagency coordinating committee (ICC) that functions in lieu of NITAG | Yes: NITAG which meets 1-3 WHO criteria | Yes: NITAG which meets 4-5 WHO criteria | Yes: NITAG which meets all 6 WHO criteria |
|  | 9) Political will pertaining to PCV introduction<sup>e</sup> | No Information | Political interest present, along with strong political challenges to vaccine introduction | Expression of interest via reports and interviews of stakeholders | - | Official statement of intent from the government |
|  | 10) Gavi-eligibility and support | Non-Gavi | MIC<sup>e</sup> approach: Never Gavi-eligible | MIC<sup>f</sup> approach: Former Gavi-eligible | Gavi-eligible | Gavi-eligible (fragile and conflict segment)<sup>g</sup> |

<sup>a</sup> The greater the score, the lower the barriers to vaccine introduction.
<sup>b</sup> Estimated incidence (2015) of pneumococcal disease including pneumonia, severe pneumonia, meningitis and invasive non pneumonia non meningitis (NPNM).
<sup>c</sup> World Bank Child Mortality Data. Available at: https://data.worldbank.org/indicator/SH.DYN.MORT
<sup>d</sup> The six process indicators for NITAGs are the presence of: (1) a legislative or administrative basis, (2) formal terms of reference, (3) five or more areas of expertise represented among its membership, (4) at least one meeting per year, (5) distribution of the agenda and background documents at least one week prior to meetings, and (6) mandatory disclosure of conflict of interests
<sup>e</sup> Sourced from government reports, public announcements, published accounts.
<sup>f</sup> Gavi's MICs Approach supports selected middle-income countries to drive sustainable introduction of key missing vaccines in select former- and never Gavi-eligible countries. [Eligible country list, July 2022 https://www.gavi.org/sites/default/files/programmes-impact/support/Countries-and-economies-eligible-for-support-under-Gavi-MICs-Approach.pdf]
<sup>g</sup> Gavi has identified a set of countries that face chronic fragility to provide more flexible and tailored support – these countries in our scoring received a score of 4, considering that Gavi will prioritize these countries with bespoke interventions to adopt within the conflict affected environment.

The ‘health characteristics’ domain included disease burden indicators suggestive of ‘need’ for PCV, and national health expenditure as a marker of health system readiness for PCV introduction. We used estimated incidence rate of pneumococcal pneumonia from 2015 as the indicator to account for the number of new cases observed in each country (22,23). A higher disease incidence rate and under five child mortality rate suggested a greater urgency for PCV introduction and were given higher scores. Countries whose health expenditure formed a significant proportion of their gross domestic product (GDP) were expected to be more supportive of PCV introduction (24). World Bank resources were used to determine country characteristics such as population, under five child mortality rate, GDP and national health expenditure (20).

The domain of ‘immunization systems’ included national coverage of childhood vaccines (DTP3 and MCV1) (25), and introduction of at least one of the four ‘new’ vaccines – RV, HPV, IPV, and MCV2. We assumed that countries that have supported the introduction of these vaccines may be more likely to have financial and health system resources as well as momentum to introduce vaccines, such as PCV (25,25–27). Within this domain, we also included an indicator for partial PCV introduction, which takes into consideration countries that have introduced PCV sub-nationally and/or to risk groups. We gave countries a higher score if they have partially introduced PCV assuming that they are closer to introducing the vaccine nationally. WHO databases were used to obtain information on vaccine coverage and introductions (28).

Within the ‘policy framework’ domain, relevant indicators included the presence of appropriate technical advisory bodies, political will, and external support from Gavi. With regards to technical advisory bodies, we considered the presence of National Immunization Technical Advisory Groups (NITAGs) or Interagency Coordinating Committees (ICCs) for each country. NITAGs are independent committees that provide recommendations to the Ministries of Health (29) which is a requirement to apply for Gavi support. According to the Global NITAG Network (GNN), a NITAG is considered functional when it meets six criteria laid out by the WHO; having a legislative or administrative basis; having formal terms of reference; having at least five areas of expertise among its membership; having at least one meeting per year; distribution of the agenda and background documents shared at least one week prior to meetings; and having mandatory disclosure of conflicts of interest (30–33). In African countries, ICCs play a similar role to NITAGs, acting as a mechanism for coordination between national Expanded Program on Immunization (EPIs) and bilateral or multilateral partners (34). To categorize degrees of political will in support of PCV introduction, we considered official government statements indicating intent to introduce PCV, in-country stakeholders’ reports available on official websites or in gray literature. Considerations for external support included Gavi-eligibility for vaccine introduction support or catalytic support to Middle Income Countries (MICs) (28), including identification of countries that face chronic fragility and those countries that fall within the Gavi 5.0 Strategy for MICs that was approved in June 2022(35). For countries that qualify under this MIC strategy, we considered two further categories in our scoring – never-Gavi eligible and former-Gavi eligible – with the assumption that countries that were formerly eligible for Gavi support are more familiar with the processes and mechanisms compared to countries that have never been exposed to Gavi support, and as such were given differential scores. While the fragility context due to conflict and other emergencies can itself be a significant barrier to new vaccine introduction, we elected to assign a higher score for this factor, considering the existing Gavi ‘Fragility, emergencies and displaced populations policy’ that prioritizes affected countries so that they can benefit from greater international attention and additional flexible and differentiated support, which can lower financial and logistical barriers to vaccine introductions (36).

### Validation of scoring framework

To verify that the domains and indicators selected for our assessment were suitable and our scoring could appropriately reflect the landscape for future PCV introduction, we performed the same domain-indicator mapping and scoring exercise with eight countries that had recently introduced PCV in 2021 and 2022 (India, Indonesia, Samoa, Tajikistan, Timor-Leste, Tonga, Tuvalu, and Vanuatu). We hypothesized that the selected indicators and scoring system may be deemed appropriate and valid if all or most of these recently introduced countries would fall within the ‘low-barrier’ country archetype.

## Results

After scoring countries based on indicators described in previous section, the total score for each country was obtained through summation of the country’s individual scores across the ten indicators. Countries were then divided into tertiles based on the highest (score of 23) and lowest (score of 12) observed (Table 3). Countries were categorized into three archetypes: those with low barriers to introduction called ‘low-barrier countries (scores 20 to 23); ‘moderate-barrier countries’ (scores 16 to 19); and ‘high barrier-countries’ (scores 12 to 15).

**Table 3:**
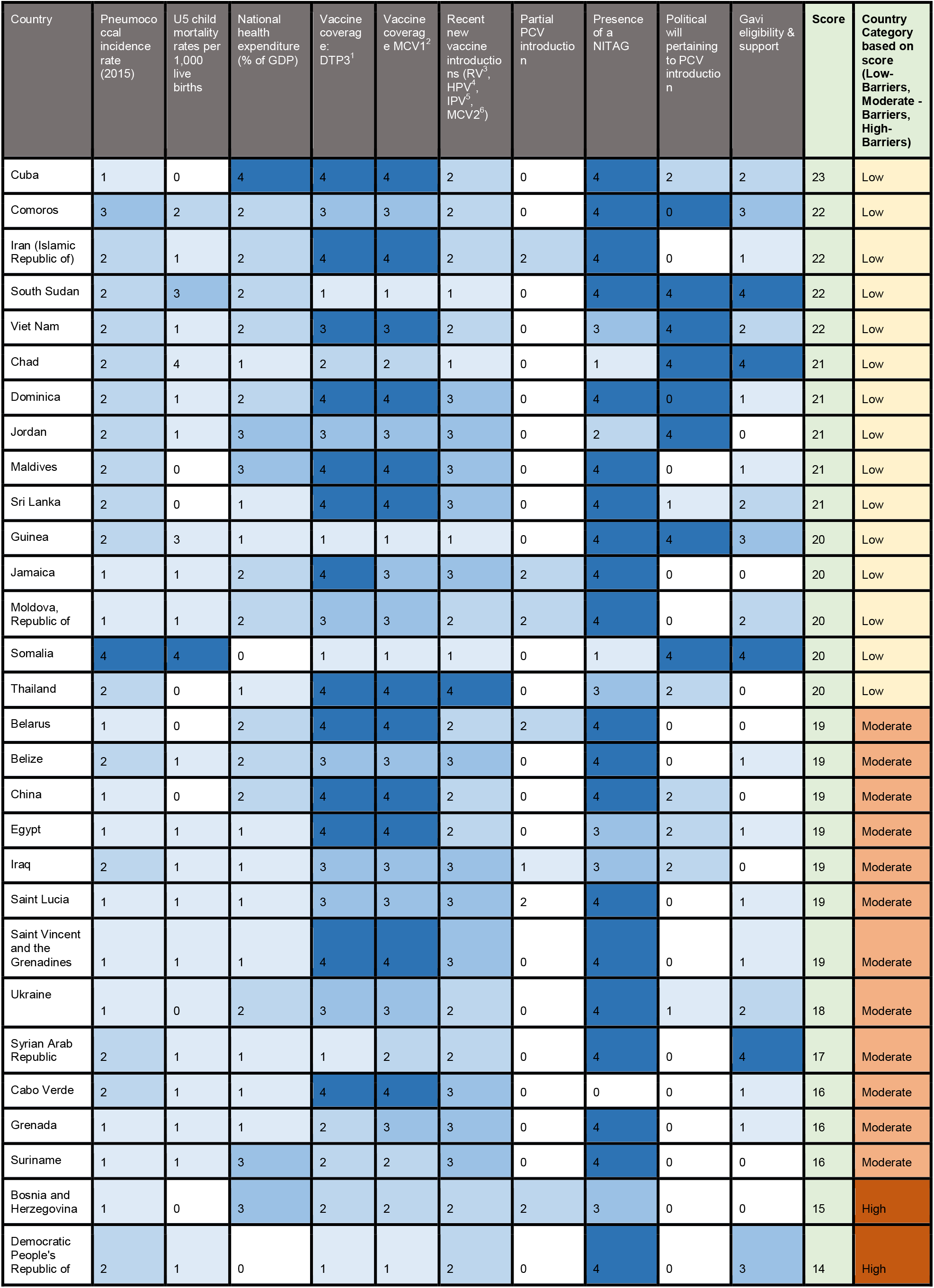
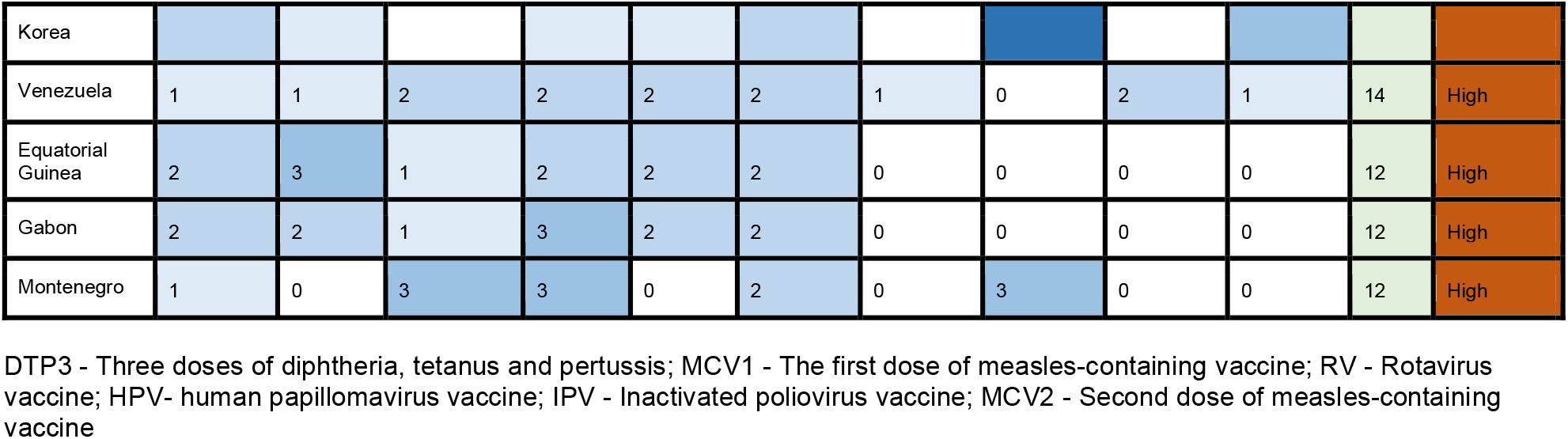
Scoring and categorization of ‘last-mile’ countries yet to introduce PCV

Based on the synthesis of the ten indicator scores, the 33 countries were assigned into three archetypes, with fifteen countries (45 %) in the ‘low barriers to introduction’ archetype, twelve countries (36 %) in the moderate barriers to introduction’ archetype, and six countries (18 %) in the ‘high barriers to introduction’ archetype (Figure 1, Table 4).

**Figure 1.**
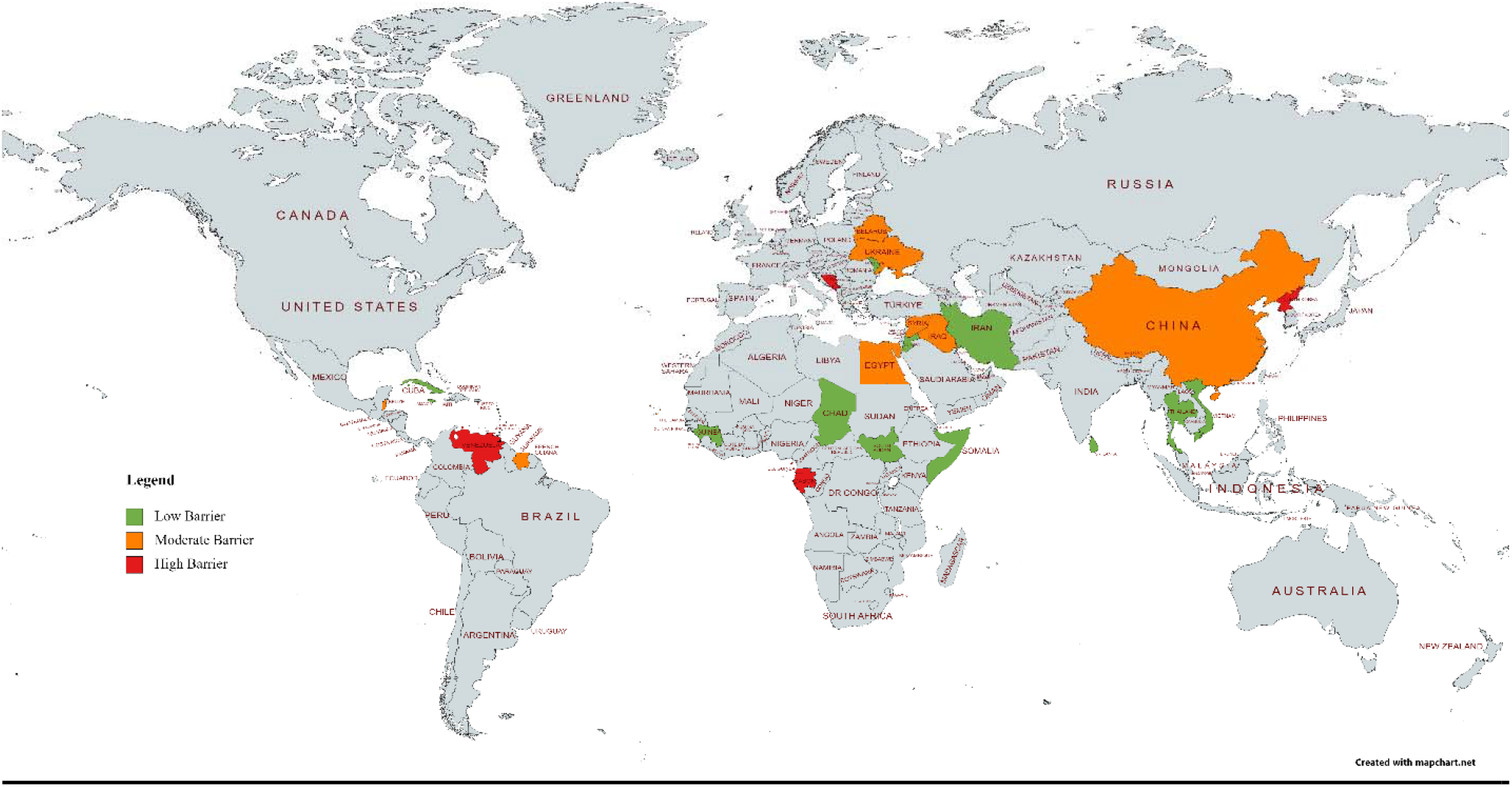
Map of 33 LMICs yet to introduce PCV with their respective archetypes ^*^Venezuela is currently unclassified by the World Bank, but for the purposes of this exercise and categorization process, it is considered as a LMIC (source: **https://datahelpdesk.worldbank.org/knowledgebase/articles/906519-world-bank-country-and-lending-groups**)

**Table 4:** LMICs that have not yet introduced PCV, categorized as per their archetypes for potential PCV introduction

| Category | Countries |
| --- | --- |
| 1. Low barriers to introduction (Score 20 to 25) | 1. Cuba<br>2. Comoros<br>3. Iran (Islamic Republic of) <sup>a</sup><br>4. South Sudan<br>5. Viet Nam<br>6. Chad<br>7. Dominica<br>8. Jordan<br>9. Maldives<br>10. Sri Lanka<br>11. Guinea<br>12. Jamaica<br>13. Moldova, Republic of<br>14. Somalia<br>15. Thailand |
| 2. Moderate barriers to introduction (Score 15 to 19) | 1. Belarus<br>2. Belize<br>3. China<br>4. Egypt<br>5. Iraq <sup>b</sup><br>6. Saint Lucia<br>7. Saint Vincent and the Grenadines<br>8. Ukraine<br>9. Syrian Arab Republic<br>10. Cabo Verde<br>11. Grenada<br>12. Suriname |
| 3. High barriers to introduction (Score 8 to 12) | 1. Bosnia and Herzegovina<br>2. Democratic People's Republic of Korea<br>3. Venezuela <sup>b</sup><br>4. Equatorial Guinea<br>5. Gabon<br>6. Montenegro |

<sup>a</sup> Countries that have partially introduced PCV among certain regions of the country and/or to specific risk groups.
<sup>b</sup> Iraq and Venezuela both previously introduced PCV in 2017 and 2014 respectively, however halted its use in the national routine schedule (19).

**Table 5:** List of countries with opportunities that lower barriers to PCV introduction

| Archetype | Gavi-eligible<br>(*Fragile and conflict<br>segment) | Gavi MICs Approach |  | Non-Gavi countries |
| --- | --- | --- | --- | --- |
|  |  | *Never Gavi-eligible* | *Former Gavi-eligible* |  |
| **Low-barrier countries** | Chad*<br>Comoros<br>Guinea<br>Somalia*<br>South Sudan* | Dominica<br>Iran (Islamic Republic of)<br>Maldives | Cuba<br>Moldova, (Republic of)<br>Sri Lanka<br>Viet Nam | Jamaica<br>Jordan<br>Thailand |
| **Moderate-barrier countries** | Syrian Arab Republic* | Belize<br>Cabo Verde<br>Egypt<br>Grenada<br>Saint Lucia<br>Saint Vincent and the Grenadines | Ukraine | Belarus<br>China<br>Iraq<br>Suriname |
| **High-barrier countries** | Democratic People's Republic of Korea | Venezuela |  | Bosnia and Herzegovina<br>Equatorial Guinea<br>Gabon<br>Montenegro |

### Low barriers to introduction country archetype

These fifteen countries have fewer barriers for introducing vaccines, such as PCV. Moreover, they also experience higher disease burden and child mortality rates signifying vaccine need. In addition, these countries have factors indicative of favorable circumstances for PCV introduction and more successful rollout, such as high immunization coverages of DTP3 and MCV2, recent new vaccine introductions, substantive or reasonable expenditure on health, eligibility for Gavi support, and a functioning NITAG.

### Moderate barriers to introduction country archetype

The 12 countries within this archetype have mixed levels of barriers; while most experience low or moderate disease burden, political will is not high for diverse reasons, and Gavi support scores are relatively low, as seven out of 12 (58%) are MICs. These mixed factors cause the overall score to fall within the moderate range, indicating greater complexity in PCV introduction. Some countries, such as Ukraine and Syrian Arab Republic, had conditions conducive to PCV introduction prior to ongoing conflict, however due to present circumstances, fall in the ‘moderate-barrier’ category (37).

### High barriers to introduction country archetype

The six countries in this archetype score in the lower range with regards to political will and health systems such as NITAG functionality or vaccine coverage, indicating a more challenging environment for PCV introduction. Validation of our scoring framework revealed that seven out of eight countries that had recently introduced PCV in 2021 and 2022 fell within the “low-barriers” archetype (See Supplementary Table S2). Only Vanuatu, which introduced PCV in 2021, fell within the “moderate barriers” archetype.

## Discussion

To achieve the global target of 90% vaccine coverage by 2030 and address persistent global vaccine access inequities and high child mortality, our archetypal analysis of the remaining ‘last-mile’ countries highlights similar yet diverse barriers across non-PCV nations. Among LMICs, 33 countries are yet to introduce PCV and are the focus of this analysis. Country groupings using diverse indicators for vaccine decision-making offer a framework to guide national and global health agencies to design tailored PCV advocacy and implementation strategies to enable national PCV introduction.

Two major cross-cutting barriers to PCV introduction are conflicting country priorities and cost of PCV. Previous studies have found that vaccine adoption was related to donor financing, co-financing uncertainty, and price per dose (38–41). PCV’s cost per dose is one of the highest among routine childhood vaccines and has presented as a major barrier to introduction in many countries (42). As of 2023, the two widely available PCV products cost up to $226.43 per dose in the U.S. private market (43). When purchased through the Pan American Health Organization (PAHO) Revolving Fund, a cooperative mechanism for joint procurement of vaccines and related supplies for PAHO countries, PCV costs USD$2-11.76 per dose (44). MICs can also access PCV through UNICEF at around $14-45 per dose, depending on the product (45). Major efforts to encourage global PCV introduction have helped bring product prices down and expand the global PCV market (46,47). The 2023 UNICEF and Gavi-related pricing for Gavi-eligible countries of PCV products in various presentations ranges from USD $1.50 (a time-bound discounted price that is normally USD$2.00) to $3.30 per dose (48,49). This lower pricing trend has created an enabling atmosphere for introduction decisions.

Gavi support plays a significant role in lowering barriers to PCV introduction. Being Gavi-eligible for financing support lowers the cost barrier for PCV. Of the countries that have not yet introduced PCV, seven (21%) are Gavi-eligible, of which, five are ‘low-barrier’ archetypes (Chad, Comoros, Guinea, Somalia and South Sudan). Four of these countries, Chad, Somalia, South Sudan, and Syrian Arab Republic are additionally eligible for flexible and tailored support from Gavi under the Fragility, Emergencies and Displaced Populations policy (36). Syrian Arab Republic and Democratic People’s Republic of Korea, while both Gavi-eligible, fall within the ‘moderate’ and ‘high’ barrier archetypes respectively. Concerted efforts to enable eligible countries leverage these opportunities would bring the world closer to achieving IA2030’s goals for PCV.

15 out of the 18 (83 %) countries classified as moderate- to high-barrier are MICs, which typically have a much slower rate of introducing new and underutilized vaccines, including PCV (50). This may be likely due to lower disease burden, lower external support, and barriers of cost and product visibility (51). Yet MICs together house an overall greater number of vulnerable populations compared to low-income countries and consequently report 67% of global vaccine-preventable deaths (52,53). Many of the world’s vulnerable populations are in MICs, where median gross national income per capita ranges widely between US$1,806 and $13,205 per year, and inequities within MICs are being exacerbated by increasing migration, urbanization, conflict, and climate change (20,51). Recognizing the unique and precarious position of MICs, calls to action have highlighted the need for country ownership, collaborative planning, and joint funding (52). Gavi’;s MICs Approach allows select MICs to request support from Gavi to address barriers to introduce vaccines, for instance through advocacy, technical assistance, and peer-to-peer learning (35). 15 non-PCV MICs currently qualify for this support, including seven ‘low barrier’ countries, seven ‘moderate barrier’ countries and one ‘high barrier’ country. This leaves 11 upper-middle-income, non-PCV countries ineligible for Gavi support under the MICs Approach, that are spread across all three archetypes, which underscores the need for innovative and tailored approaches for advocacy for countries with diverse barriers and lack of packaged opportunities. As an example of a collaborative advocacy approach for a country that was ineligible for specific funding flexibilities, Jordan’s NITAG was provided with evidence synthesis on PCV safety, efficacy, product and cost options, leading to a recent NITAG recommendation for PCV introduction using repurposed COVID-19 vaccine funds (54).

The ‘low-barrier’ archetype countries are well positioned in terms of health system strength and policy support for PCV introduction. These countries require nuanced strategies and have unique strengths and weaknesses for their individual country settings in their ability to introduce PCV. A call to action to enable high-burden, low-barrier countries with high political will, such as Chad, Guinea, Somalia, and South Sudan, can be a powerful approach to bring together and garner support from diverse national and international agencies (55). On the other hand, tailored strategies are required for some countries, for example Sri Lanka scores as a low-barrier country showing favorable indicators for introduction, including being eligible for Gavi support under the MICs Approach, however is experiencing a severe economic crisis that constrains political will for vaccine introduction (56).

Within our scheme, countries with a higher disease burden are more likely to be classified as having a low barrier to introduction yet may require introduction support to achieve public health impact. Studies across high-burden African settings (The Gambia, South Africa, Kenya, Rwanda, Burkina Faso, and Zambia) have shown that PCV introduction has significantly reduced cases, hospitalizations, and deaths associated with pneumonia (57–62). PCV introduction in the remaining high-burden countries is essential for saving child lives (50).

Modeled estimates suggest that routinely vaccinating children between 0-1 years of age with PCV between 2000 and 2030 could avert approximately 32,600 child deaths in Somalia, 11,300 child deaths in Guinea, 11,900 child deaths in South Sudan, and 35,000 child deaths in Chad (7,63). With mechanisms such as Gavi’s co-financing support, UNICEF procurement and reduced prices for low-income countries, as well as tailored support for countries facing chronic fragility, there is continued support from the global community to facilitate the introduction of vaccines in those countries with the greatest need.

The ‘moderate-barrier’ archetype countries were diverse in their PCV-introduction barriers. Egypt shows a strong immunization system, although competing country priorities and cost barriers preclude imminent PCV introduction. China has the capacity to locally manufacture new PCV products, however, ineligibility for Gavi support could also prolong introduction while waiting for the licenses and logistics for their new product (64,65). All 11 countries within the moderate-barrier archetype are MICs. As such, enhanced international coordination and advocacy for greater flexibility and transparency on product pricing is warranted. Additionally, stronger procurement mechanisms and strengthened in-country and regional partnerships could help these countries in enabling PCV introduction. Examples of countries that would have otherwise made progress with respect to PCV introduction are Ukraine and Syrian Arab Republic, where efforts have stalled due to their ongoing geopolitical situation indicating how deeply conflict can set back health system progress.

The ‘high-barrier’ archetype countries have lower or unstated political will with respect to PCV introduction compared to the moderate and lower barrier archetypes, which may be linked with lower disease burden and lower relative health spending. Lower levels of engagement and communication with the international community along with political instability and conflict status may also explain some of these barriers (Bosnia and Herzegovina, Democratic People’s Republic of Korea, and Venezuela).

Recent childhood vaccine introductions as a marker of health system resourcefulness are an indicator we applied to signal the likelihood of introducing PCV. While 127 countries introduced COVID-19 vaccines in 2021, many fear that the momentum to introduce other vaccines has decreased, partially due to pandemic-related health system fatigue (10). On the other hand, some countries have elected to introduce both PCV and RV concurrently, leveraging the co-introduction strategy to streamline implementation activities; this can increase efficiency and may generate substantial reductions in respiratory and diarrheal infections within a shorter period of time (26,66). Three countries in our analysis have already introduced RV, all of which two fall into the ‘low-barrier’ archetype (Jordan, Moldova and Thailand).

Vaccine confidence among community members and policy makers can be a facilitator or barrier in country vaccine introduction decisions. Besides being uncertain and dynamic, vaccine confidence is challenging to quantify, and hence was not included in our set of indicators. Community-level vaccine hesitancy concerns have been well-documented in the literature, particularly hesitancy surrounding COVID-19 vaccines (67). Concerns driving vaccine hesitancy have traditionally included fears of side effects, spread of misinformation, lack of provider recommendation, and reduced belief that vaccines are valuable (67–69). Studies have provided limited insights into whether hesitancy among decision-makers may impact decisions to facilitate vaccine introduction, and warrants further qualitative research. Related insights could inform tailored communication and advocacy strategies that encourage PCV introduction and uptake.

Several study constraints exist. First, there is subjectivity that is inherent in archetyping, which we tried to reduce through a quantitative scoring approach and selected indicators. These indicators were diverse in nature and included ‘need-related’ parameters such as disease burden as well as ‘modifiable constructs’ such as National Immunization Systems and political will. The ‘Gavi-eligibility and support’ indicator takes into consideration the additional support that some countries receive on account of fragility, conflict, or emergencies – resulting in a higher score, although the fragility aspect may intrinsically increase barriers to vaccine introduction. Second, data may be limited in quality and availability as information on countries lacking accessible health ministry websites may have been missed or absent. Pricing data for individual countries were not always available (39). Searches were conducted in English, which may have also limited the results. Information regarding national political will is inconsistently available, and trajectories for PCV introduction may have changed since the writing of this manuscript. Finally, as this is a desk landscaping exercise, results were not validated with surveys of interviews of in-country stakeholders. Heterogeneous article types were included as sources, including gray literature and proceedings of consultations and meetings, which are not peer reviewed and may be biased. Nevertheless, our archetypal analysis provides a reasonable framework to guide advocacy strategies for PCV vaccine introduction, especially in the 33 LMICs yet to do so.

Recommendations can be based by archetypes, as identification of similar barriers within country archetypes point to a way forward to accelerate PCV introduction in these last-mile countries. Select countries within the ‘low barrier’ archetype and those that are eligible for Gavi support could benefit from timely assistance in Gavi application completion, as increased international support pathways through existing schemes together with in-country commitment can enable PCV rollout in the near term. Notwithstanding the diversity among countries within the ‘moderate barriers to introduction’ archetype, current approaches could focus on building resilient systems, NITAG strengthening, vaccine delivery enhancement, and appropriate vaccine-related public health messaging. For countries within the ‘high barriers to introduction’ archetype, greater engagement and inclusivity within the international community could be a strategy to lower barriers, together with increased global engagement for health system strengthening and enhanced decision-making support for new vaccine introduction. While these recommendations are targeted towards each of the three archetypes, several of these approaches may be relevant for all non-PCV countries. With only half the world’s children receiving PCV as of 2021 (9), it is clear that investing in new approaches to strengthen financing and country primary healthcare systems, and leveraging scientific and technological innovations are urgently needed, as articulated by UNICEF’s The State of the World’s Children 2023 that was fully dedicated to vaccination (69).

## Conclusion

This study is a thorough assessment of a variety of factors within countries that inform strategies for PCV introduction. These results provide a reasonable and actionable framework that may be used by the global community to recognize and address distinct barriers to PCV use in non-PCV last-mile countries. Implementation approaches that emerge from this framework are responsive to the global call to action for increasing global PCV coverage. Closing the PCV gap will entail concerted and robust responses in increasing political, scientific, logistical, and financial ownership by countries and global agencies to enhance child survival worlwide.

## Data Availability

All data produced in the present study are available upon reasonable request to the authors.

https://data.worldbank.org/indicator/

https://immunizationdata.who.int/index.html

## Supplementary Materials

**Table S1:**
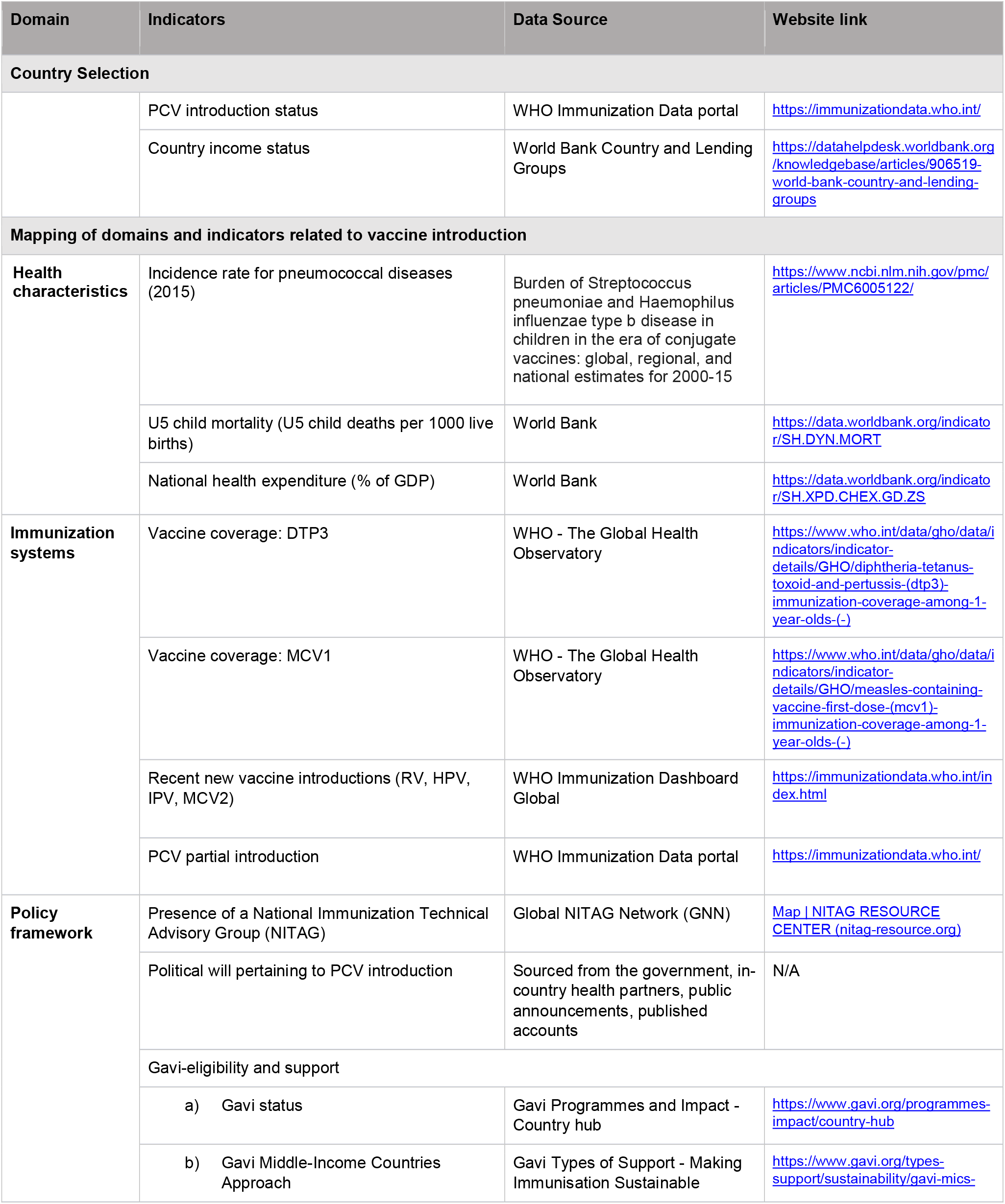
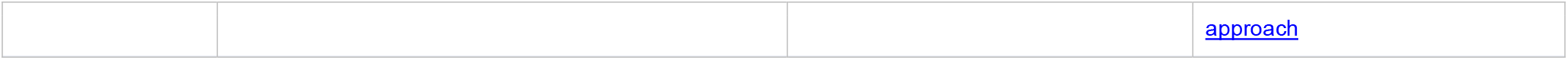
Data Sources (website links and references) for country selection, landscape desk review and domain-indicator mapping

| Domain | Indicators | Data Source | Website link |
| --- | --- | --- | --- |
| **Country Selection** |  |  |  |
|  | PCV introduction status | WHO Immunization Data portal | https://immunizationdata.who.int/ |
|  | Country income status | World Bank Country and Lending Groups | https://datahelpdesk.worldbank.org/knowledgebase/articles/906519-world-bank-country-and-lending-groups |
| **Mapping of domains and indicators related to vaccine introduction** |  |  |  |
| **Health characteristics** | Incidence rate for pneumococcal diseases (2015) | Burden of Streptococcus pneumoniae and Haemophilus influenzae type b disease in children in the era of conjugate vaccines: global, regional, and national estimates for 2000-15 | https://www.ncbi.nlm.nih.gov/pmc/articles/PMC6005122/ |
|  | U5 child mortality (U5 child deaths per 1000 live births) | World Bank | https://data.worldbank.org/indicator/SH.DYN.MORT |
|  | National health expenditure (% of GDP) | World Bank | https://data.worldbank.org/indicator/SH.XPD.CHEX.GD.ZS |
| **Immunization systems** | Vaccine coverage: DTP3 | WHO - The Global Health Observatory | [https://www.who.int/data/gho/data/indicators/indicator-details/GHO/diphtheria-tetanus-toxoid-and-pertussis-(dtp3)-immunization-coverage-among-1-year-olds-(-)](https://www.who.int/data/gho/data/indicators/indicator-details/GHO/diphtheria-tetanus-toxoid-and-pertussis-%28dtp3%29-immunization-coverage-among-1-year-olds-%28-%29) |
|  | Vaccine coverage: MCV1 | WHO - The Global Health Observatory | [https://www.who.int/data/gho/data/indicators/indicator-details/GHO/measles-containing-vaccine-first-dose-(mcv1)-immunization-coverage-among-1-year-olds-(-)](https://www.who.int/data/gho/data/indicators/indicator-details/GHO/measles-containing-vaccine-first-dose-%28mcv1%29-immunization-coverage-among-1-year-olds-%28-%29) |
|  | Recent new vaccine introductions (RV, HPV, IPV, MCV2) | WHO Immunization Dashboard Global | https://immunizationdata.who.int/index.html |
|  | PCV partial introduction | WHO Immunization Data portal | https://immunizationdata.who.int/ |
| **Policy framework** | Presence of a National Immunization Technical Advisory Group (NITAG) | Global NITAG Network (GNN) | [Map \| NITAG RESOURCE CENTER (nitag-resource.org)](https://nitag-resource.org/) |
|  | Political will pertaining to PCV introduction | Sourced from the government, in-country health partners, public announcements, published accounts | N/A |
|  | Gavi-eligibility and support |  |  |
|  | a) Gavi status | Gavi Programmes and Impact - Country hub | https://www.gavi.org/programmes-impact/country-hub |
|  | b) Gavi Middle-Income Countries Approach | Gavi Types of Support - Making Immunisation Sustainable | https://www.gavi.org/types-support/sustainability/gavi-mics- |

|  |  |  |
| --- | --- | --- |
|  |  | approach |

**Table S2:**
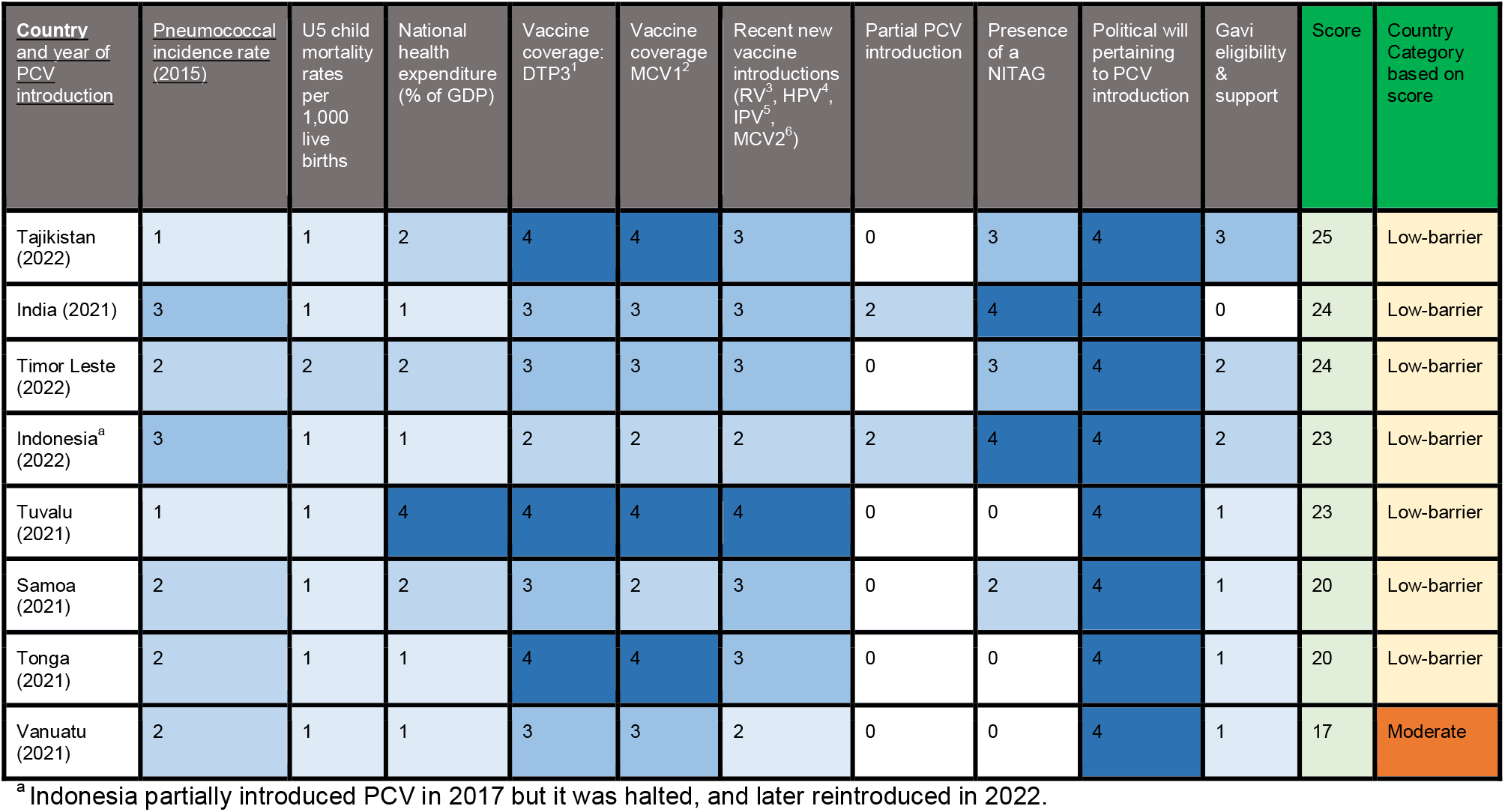
Validation of scoring framework, countries that introduced PCV in 2021 and 2022

